# Scoring Physician Risk Communication in Prostate Cancer Using Large Language Models

**DOI:** 10.1101/2025.08.07.25333034

**Authors:** Guillermo Lopez-Garcia, Dongfang Xu, Michael Luu, Renning Zheng, Timothy J. Daskivich, Graciela Gonzalez-Hernandez

**Affiliations:** Department of Computational Biomedicine, Cedars-Sinai Medical Center, Los Angeles, CA, USA; Department of Biostatistics, Cedars-Sinai Medical Center, Los Angeles, CA, USA; Department of Urology, Cedars-Sinai Medical Center, Los Angeles, CA, USA; Tsinghua Medicine, Tsinghua University, Beijing, China

**Keywords:** Prostate cancer, physician-patient communication, risk communication, artificial intelligence, large language models, natural language processing, shared decision-making

## Abstract

Effective risk communication is essential to shared decision-making in prostate cancer care. However, the quality of physician communication of key tradeoffs varies widely in real-world consultations. Manual evaluation of communication is labor-intensive and not scalable. We present a structured, rubric-based framework that uses large language models (LLMs) to automatically score the quality of risk communication in prostate cancer consultations. Using transcripts from 20 clinical visits, we curated and annotated 487 physician-spoken sentences that referenced five decision-making domains: cancer prognosis, life expectancy, and three treatment side effects (erectile dysfunction, incontinence, and irritative urinary symptoms). Each sentence was assigned a score from 0 to 5 based on the precision and patient-specificity of communicated risk, using a validated scoring rubric. We modeled this task as five multiclass classification problems and evaluated both fine-tuned transformer baselines and GPT-4o with rubric-based and chain-of-thought (CoT) prompting. Our best performing approach, which combined rubric-based CoT prompting with few-shot learning, achieved micro averaged F1 scores between 85.0 and 92.0 across domains, outperforming supervised baselines and matching inter-annotator agreement. These findings establish a scalable foundation for AI-driven evaluation of physician–patient communication in oncology and beyond.

## 1. Introduction

Effective communication of risk during treatment decision-making is essential for achieving high-quality, patient-centered care in oncology.^1,2^ In prostate cancer, where multiple treatment options exist with similar oncologic outcomes but differing side-effect profiles, shared decisionmaking (SDM) is particularly critical.^3,4^ To support this process, the American Urological Association (AUA) recommends that clinicians address five core domains during consultations: cancer severity, life expectancy (LE), cancer prognosis (CP), baseline function, and treatment side effects.^5^ Among these, life expectancy,^6,7^ cancer prognosis, ^8,9^ and side effects represent the most critical tradeoffs patients must consider when evaluating treatment options.^10^

Despite the published guidelines, physician communication around these key concepts remains highly variable in both frequency and quality.^11^ Recent analyses of prostate cancer consultations revealed that life expectancy, cancer prognosis, and side effects are often omitted entirely or discussed without sufficient quantification.^11,12^ Even when mentioned, the level of detail ranges from general remarks to highly tailored, patient-specific risk estimates. This inconsistency poses challenges to effective shared decision-making, particularly given that many patients express a preference for quantified, personalized risk information.^7^ Such variability not only undermines patient understanding but may contribute to poor decision quality and future regret,^12^ highlighting the need for scalable tools to systematically assess the quality of physician risk communication.

While the value of high-quality risk communication is well recognized, practical methods for evaluating how effectively physicians communicate these tradeoffs are limited in real-world clinical settings.^13^ Manual transcript coding remains the predominant approach for assessing communication, but this process is labor-intensive, costly, and not feasible at scale.^14^ Recent advances in natural language processing (NLP)^15,16^ offer new opportunities to automate these evaluations at scale, with greater efficiency and consistency.

Our recently published study^17^ was the first to leverage NLP to evaluate physician communication quality in prostate cancer consultations. In that work, we developed supervised machine learning models to accurately retrieve sentences pertaining to five central decisionmaking domains—cancer severity, life expectancy, cancer prognosis, baseline function, and treatment side effects. We demonstrated that identifying the presence and frequency of such domain-specific content could serve as a proxy for broader consultation quality. However, the focus of that approach was limited to detecting whether key topics were discussed, without evaluating the depth, precision, or patient-specificity of how risks were communicated. In contrast, the present study moves beyond content retrieval to introduce a structured, rubric-based assessment of communication quality for each sentence. By assigning fine-grained quality scores that reflect not just topic presence but also the clarity, specificity, and quantitative precision of risk information, our framework offers a substantially more nuanced and actionable evaluation of physician–patient communication in prostate cancer care.

We introduce a novel framework for sentence-level scoring of communication quality using large language models (LLMs). Rather than merely detecting relevant content, we assess the precision and patient-specificity of each sentence using a validated scoring rubric.^14^ Five core communication tradeoffs central to shared decision making in prostate cancer are assessed: cancer progression (CP), life expectancy (LE), and three common treatment side effects: erectile dysfunction (ED), incontinence (INC), and irritative urinary symptoms (IUS). We formulate the task as five multiclass classification problems, one per tradeoff, where each sentence is assigned a numeric score (0–5) based on communication quality corresponding to the rubric.^14^ To address this complex task, we evaluate two prompting strategies: rubric-only prompting and rubric-based chain-of-thought (CoT) prompting,^18^ both in a few-shot in-context learning setting.^19^

By assigning quality scores grounded in established rubrics, we are able to capture not only whether key domains are discussed, but also the degree to which information is tailored, quantified, and relevant to individual patients. This enables systematic differentiation between vague, general statements and precise, patient-specific risk estimates—an essential distinction for understanding and improving shared decision-making. Leveraging the reasoning capabilities of advanced LLMs, our experiments demonstrate that rubric-informed assessment, particularly when combined with explicit chain-of-thought reasoning, allows models to approach expert-level performance even with limited annotated data.

Our contributions are threefold: (1) we develop a structured and reproducible annotation framework for scoring risk communication in prostate cancer based on previously validated rubrics;^14^ (2) we demonstrate the effectiveness of GPT-4o in performing this scoring task using rubric-aligned CoT prompting; and (3) we compare LLM-based performance to supervised baselines and show that LLMs can achieve expert-level agreement, even with limited training data. Ultimately, this automated sentence-level assessment establishes a scalable foundation for evaluating and enhancing physician–patient communication quality in prostate cancer care and lays the groundwork for applications in a broad range of clinical settings.

## 2. Materials and Methods

### 2.1. Data collection

We collected the transcripts of 20 physician-patient encounters of men with newly diagnosed clinically localized prostate cancer. From each encounter, we extracted only the physician-spoken content and segmented it into individual sentences. Using our recently developed NLP-based model,^13,17^ we automatically identified the top five sentences per consultation that were most likely to contain relevant content for each of the five key tradeoffs relevant to SDM: CP, LE, ED, INC, and IUS. This yielded an initial set of 100 candidate sentences for each tradeoff. We then manually reviewed and removed sentences that were incorrectly identified by the model as relevant. After filtering, we retained 89 sentences for CP, 100 for LE, 99 for ED, 99 for INC, and 100 for IUS. To enrich the contextual information surrounding each selected sentence, we expanded it into a longer text window by combining three sentences before and after the selected sentence from the original transcript.

### 2.2. Data Annotation

To ensure high-quality data for developing and evaluating our scoring system, we created a structured annotation framework grounded in our previously validated methodology for assessing the quality of risk communication related to cancer prognosis, life expectancy, and treatment side effects for prostate cancer.^14^ Our objective was to quantify the level of precision with which physicians conveyed risk-related information across the five key tradeoffs in each selected sentence.

Each text fragment was assigned a numeric score from 0 to 5, with higher values indicating more patient-specific and quantitatively precise communication. Scores were assigned separately for each selected sentence of the five tradeoffs.

Table 1 summarizes the scoring rubrics. For cancer prognosis, scores reflected the extent to which physicians quantified the risk of cancer-specific mortality, progression, or metastasis. Life expectancy scoring captured how physicians estimated patient longevity outside of their cancer diagnosis. Finally, the three treatment side effects—erectile dysfunction, urinary incontinence, and irritative urinary symptoms—shared a common rubric, focused on the specificity and clarity of communicated risk related to each outcome.

**Table 1.** Scoring rubric for Cancer Prognosis, Life Expectancy, and Side Effects. The rubrics of *Side Effects* column apply to ED, UI, and IUS.

| Score | Cancer Prognosis | Life Expectancy | Side Effects |
| --- | --- | --- | --- |
| 0 | Not mentioned | Not mentioned | Not mentioned |
| 1 | Mention without quantification (e.g., general risk mentioned) | Mention without estimation | Name only (e.g., “erectile dysfunction”) |
| 2 | Qualitative estimate only (e.g., “low risk”) | Qualitative estimate only (e.g., “long life expectancy”) | Qualitative terms only (e.g., “unlikely to affect you”) |
| 3 | Numeric estimate without treatment comparison | Rough number of years (e.g., “20–30 years”) | Numeric estimate without timeline (e.g., “30% will experience it”) |
| 4 | Numeric estimate including risk with and without treatment | Probability of survival/mortality at a timepoint | Numeric estimate with timeline (e.g., “10% at 1 year”) |
| 5 | Patient-specific numeric estimate at life expectancy, with and without treatment | Specific estimate based on patient factors (e.g., age, health status) | Patient-specific numeric estimate with timeline and reference to individual characteristics |

Two annotators with expertise in oncology and prior experience in prostate cancer communication research independently annotated all text fragments using the defined scoring rubrics for each tradeoff. The annotation guidelines also included example text fragments to illustrate each score level. Annotations were performed using Microsoft Excel, which facilitated label entry and identification of disagreements. Discrepant labels were automatically detected by comparing the scores assigned by each annotator. All disagreements were reviewed in dedicated adjudication sessions, during which annotators discussed each case and reached consensus by referring to both the rubric criteria and the corresponding text fragments.

Table 2 presents the distribution of final annotated scores across the five communication tradeoffs. Prior to adjudication, inter-annotator agreement (IAA), computed as the micro-averaged *F*_1_ score, was *F*_1_ = 85.39 for CP, *F*_1_ = 83.00 for LE, *F*_1_ = 83.84 for ED, *F*_1_ = 84.85 for

**Table 2.** Distribution of annotated quality scores (0–5) across the five communication tradeoffs. Counts and proportions (%) are shown for each score.

| Score | CP |  | LE |  | ED |  | INC |  | IUS |  |
| --- | --- | --- | --- | --- | --- | --- | --- | --- | --- | --- |
|  | Count | % | Count | % | Count | % | Count | % | Count | % |
| 0 | 22 | 24.72% | 32 | 32.00% | 14 | 14.14% | 23 | 23.23% | 65 | 65.00% |
| 1 | 2 | 2.25% | 16 | 16.00% | 28 | 28.28% | 22 | 22.22% | 13 | 13.00% |
| 2 | 3 | 3.37% | 5 | 5.00% | 11 | 11.11% | 18 | 18.18% | 2 | 2.00% |
| 3 | 25 | 28.09% | 14 | 14.00% | 16 | 16.16% | 2 | 2.02% | 6 | 6.00% |
| 4 | 27 | 30.34% | – | – | 21 | 21.21% | 30 | 30.30% | 11 | 11.00% |
| 5 | 10 | 11.24% | 33 | 33.00% | 9 | 9.09% | 4 | 4.04% | 3 | 3.00% |

### 2.3. Approach

In recent years, LLMs have demonstrated state-of-the-art performance across a wide range of NLP tasks, including classification, summarization, and reasoning tasks, without the need for supervised fine-tuning methods.^21^ These models can be adapted to new tasks using *in-context learning* (ICL) techniques, where task-specific behavior is induced through natural language instructions and a small number of labeled examples embedded in the prompt (known as “few-shot prompting”^19^). Unlike traditional supervised learning, this approach does not require parameter updates or large amounts of labeled data, making it highly adaptable for low-resource or rapidly evolving domains.^22^

For our purpose, we harness the predictive capabilities of LLMs within a few-shot ICL framework for automatically scoring physician communication about key tradeoffs in prostate cancer. We formulate the problem as five independent multiclass text classification tasks, one for each shared decision-making (SDM) tradeoff: cancer prognosis (CP), life expectancy (LE), erectile dysfunction (ED), urinary incontinence (UI), and irritative urinary symptoms (IUS). Given a text fragment from a prostate cancer consultation, the goal for each task is to assign a numeric score from 0 to 5, indicating the precision and patient-specificity with which the corresponding tradeoff is discussed. We deployed two prompting strategies for automatic scoring: (1) a rubric-based strategy, where the model is expected to follow a set of explicit scoring rules; and (2) a rubric + chain-of-thought (CoT)^18^ strategy, which further decomposes the scoring task into a sequence of structured reasoning steps. We detailed each strategy next.

#### 2.3.1. Rubric-based Prompting

In the rubric-based strategy, we constructed prompts that directly instructed the model to assign a score based on the rubric definitions (see Section 2.2). Each prompt included task instructions, the full scoring rubric for the target communication tradeoff, and one or more few-shot examples consisting of a consultation fragment and its corresponding numeric score. The output format required the model to return only a numeric score from 0 to 5. An example of the rubric-based prompt used for scoring the cancer prognosis tradeoff is shown in Table 3. The same structure was used to create prompts for the other four tradeoffs, each time substituting in the appropriate rubric and task-specific few-shot examples.

**Table 3.**
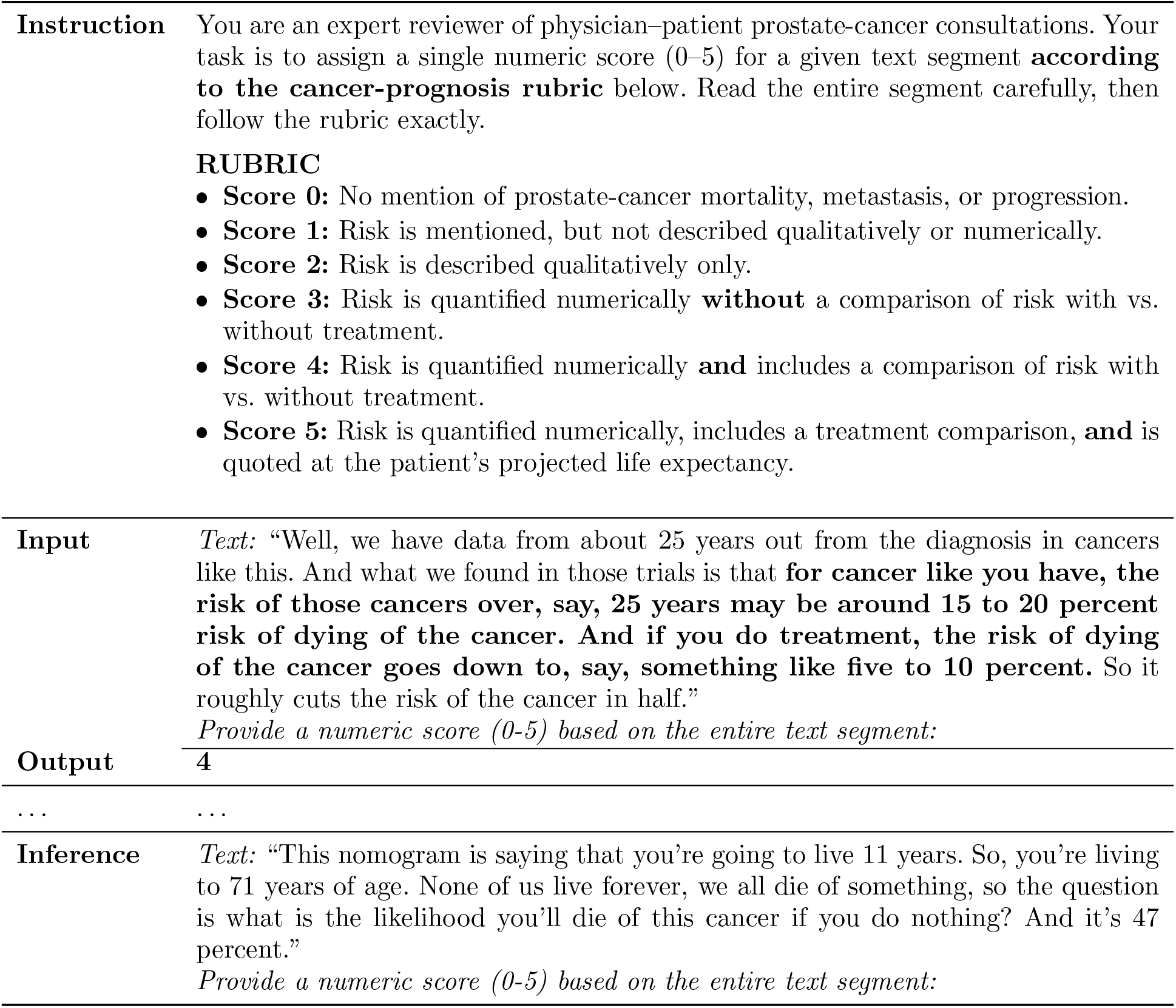
Example of rubric-based prompting for scoring the cancer prognosis tradeoff. The prompt begins with task instructions and the scoring rubric, followed by a series of few-shot examples, each comprising a text fragment from a physician–patient consultation and its corresponding numeric score.

|  |  |
| --- | --- |
| **Instruction** | You are an expert reviewer of physician–patient prostate-cancer consultations. Your task is to assign a single numeric score (0–5) for a given text segment **according to the cancer-prognosis rubric** below. Read the entire segment carefully, then follow the rubric exactly.<br>**RUBRIC**<br>• **Score 0:** No mention of prostate-cancer mortality, metastasis, or progression.<br>• **Score 1:** Risk is mentioned, but not described qualitatively or numerically.<br>• **Score 2:** Risk is described qualitatively only.<br>• **Score 3:** Risk is quantified numerically **without** a comparison of risk with vs. without treatment.<br>• **Score 4:** Risk is quantified numerically **and** includes a comparison of risk with vs. without treatment.<br>• **Score 5:** Risk is quantified numerically, includes a treatment comparison, **and** is quoted at the patient’s projected life expectancy. |
| **Input** | *Text:* “Well, we have data from about 25 years out from the diagnosis in cancers like this. And what we found in those trials is that **for cancer like you have, the risk of those cancers over, say, 25 years may be around 15 to 20 percent risk of dying of the cancer. And if you do treatment, the risk of dying of the cancer goes down to, say, something like five to 10 percent.** So it roughly cuts the risk of the cancer in half.”<br>*Provide a numeric score (0-5) based on the entire text segment:* |
| **Output** | 4 |
| ... | ... |
| **Inference** | *Text:* “This nomogram is saying that you’re going to live 11 years. So, you’re living to 71 years of age. None of us live forever, we all die of something, so the question is what is the likelihood you’ll die of this cancer if you do nothing? And it’s 47 percent.”<br>*Provide a numeric score (0-5) based on the entire text segment:* |

#### 2.3.2. Rubric + Chain-of-Thought Prompting

To encourage more interpretable and structured predictions, we also implemented a rubric + CoT prompting strategy. Chain-of-thought prompting is a technique that guides LLMs to reason through problems in a step-by-step manner, rather than producing an answer directly from the input.^18^ This approach has been shown to improve performance in tasks that require multi-step reasoning, by explicitly decomposing complex decision-making into intermediate steps.^22^ Here, we leverage CoT to augment the standard rubric-based prompt with an explicit set of reasoning instructions corresponding to each decision criterion in the scoring rubric. The model was instructed to reason through a series of decision steps that aligned with the structure of the 0–5 scoring rubric (see Table 1). Each step represented a condition that must be met to proceed to the next score level. The output format consisted of intermediate reasoning steps followed by the final score prediction. An example of the rubric + CoT prompt is shown in Table 4. The same structure was used for all five SDM tradeoffs, with each prompt adapted to reflect the decision criteria and rubric levels defined for that specific communication tradeoff.

**Table 4.**
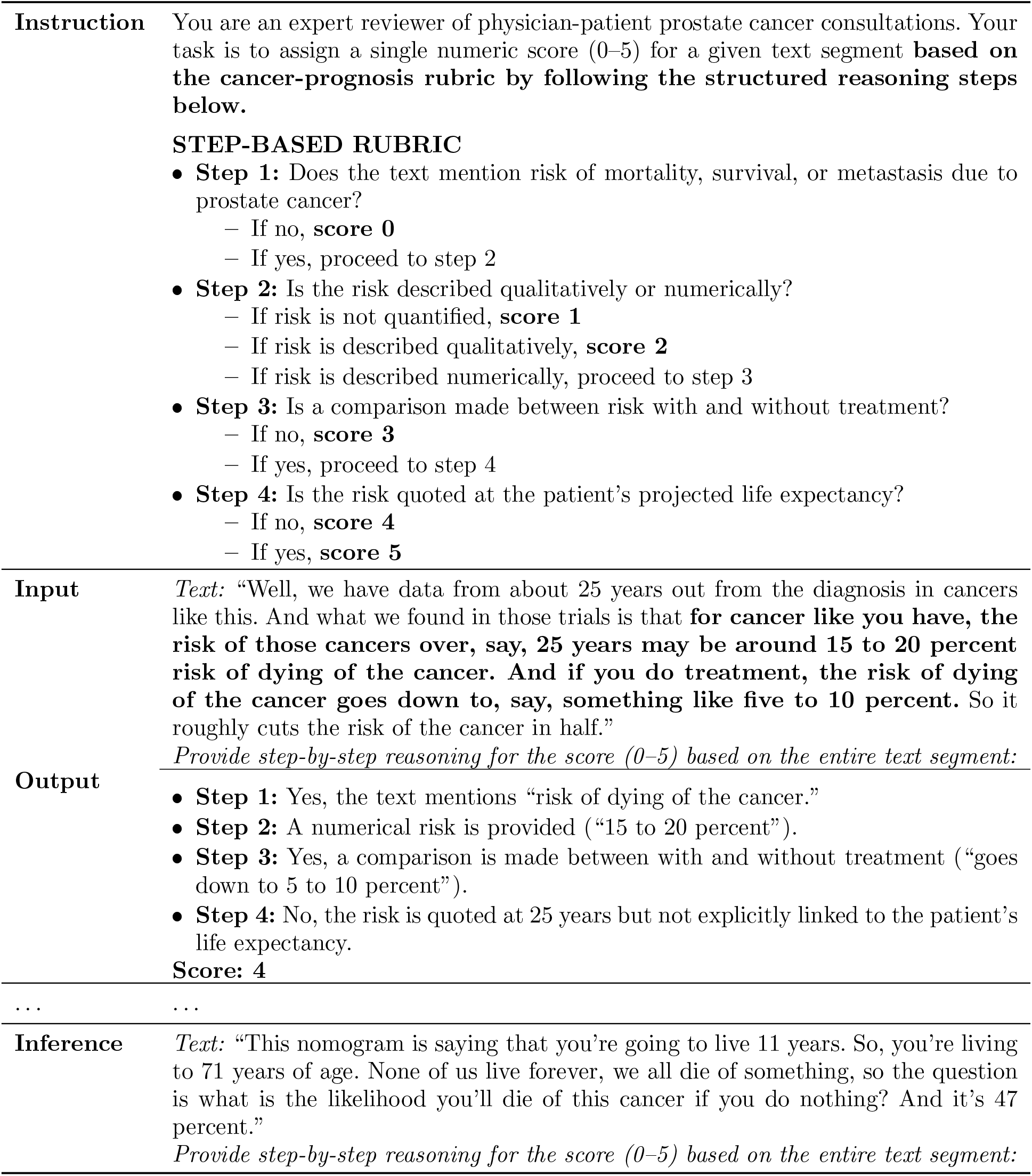
Example of rubric + chain-of-thought prompting for scoring the cancer prognosis tradeoff. Reasoning steps are required to justify the final numeric score.

### 2.4. Baseline

To evaluate the performance of our LLM-based method, we compared it against a conventional supervised learning baseline using Bidirectional Encoder Representations from Transformers (BERT)-based models.^23^ Unlike large autoregressive models such as GPT,^19^ which are optimized for open-ended generation tasks, our baseline models are encoder-based transformers trained using a masked language modeling objective.^24^ These models are more efficient to fine-tune and remain widely used for classification tasks in both general and domain-specific NLP applications. In this work, we fine-tuned BERT-based models within a multiclass text classification framework to predict physician communication scores across the five SDM tradeoffs. Specifically, we evaluated the following two model variants:

**RoBERTa-large** A large-scale variant of the RoBERTa model, pretrained on 160 GB of general-domain text using a masked language modeling objective.^25^ With approximately 355 million parameters, it removes BERT’s next-sentence prediction objective and incorporates dynamic masking to improve contextual representations.

**PubMedBERT-large** A domain-specific transformer model with 335 million parameters, pretrained from scratch on PubMed abstracts.^26^ Designed for biomedical NLP tasks, PubMedBERT is better suited to capture domain-specific terminology than general-domain models.

### 2.5. Experiments

We split each annotated dataset into training and testing sets using stratified sampling to preserve the distribution of scores (0–5). For each tradeoff, 40% of the data was used for training and 60% for testing. Specifically, the number of examples used for training/testing per tradeoff was: CP (35/54), LE (40/60), ED (39/60), INC (39/60), and IUS (40/60).

To construct the input text fragments used for both LLM prompting and baseline classifiers, we expanded each selected sentence by including the three preceding and three following sentences from the original consultation transcript. This 7-sentence context window was chosen based on prior experimentation, where we systematically compared different window sizes and found that including three sentences before and after led to the best scoring performance across tradeoffs.

Both prompting strategies were implemented using the GPT-4o model, accessed via the Microsoft Azure OpenAI API. We set the generation *temperature* to 0.3 and the *top-p* sampling parameter to 0.4, with other settings left at default values. We evaluated both few-shot and zero-shot variants of each prompting strategy:

**Zero-shot prompting**. The model received only task instructions, with no examples included in the prompt.

**Few-shot prompting**. Prompts included a small number of annotated examples spanning the full 0–5 score range. The number of examples per tradeoff (6–10) was empirically selected based on training performance, with no consistent gains observed beyond these values: 10 for CP, 7 for LE, 7 for ED, 8 for INC, and 6 for IUS.

For baseline comparisons, we trained RoBERTa-large and PubMedBERT-large classifiers on the same 40% training splits. For each tradeoff, we fine-tuned the models as 6-class (0–5 scores) classifiers using cross-entropy loss. Input text fragments were tokenized and padded to a maximum sequence length of 512 tokens. Models were trained for 10 epochs using a batch size of 6 and learning rate of 2 *×* 10^*−*5^.

### 2.6. Evaluation Metrics

We evaluated the multiclass classification performance of the baseline models and generative approaches using standard metrics: Precision, Recall, and F_1_-score. For each class label (scores 0 through 5), we define true positives (TP) as the number of fragments correctly classified with a given score, false positives (FP) as the number of fragments incorrectly predicted to have that score, and false negatives (FN) as the number of fragments with that score in the ground truth but misclassified by the model. *Precision* (P) measures the proportion of correct predictions for a given score among all instances predicted with that score 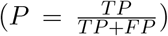; *Recall* (R) measures the proportion of correct predictions for a given score among all true instances of that score 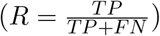; and the *F*_1_*-score* (F1) is the harmonic mean of precision and recall, balancing the trade-off between the two metrics 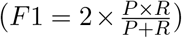. We evaluated each SDM tradeoff independently and report the micro-averaged F_1_ score to aggregate performance across all classes for each tradeoff.

## 3. Results

Table 5 presents the micro-averaged F_1_ scores for both baseline classifiers and LLM-based prompting strategies across the five SDM tradeoffs. The results show that our best-performing method (rubric + CoT with few-shot prompting) consistently outperforms both baseline classifiers and alternative prompting strategies. Specifically, this approach achieved the highest F_1_ scores across all five tradeoffs: cancer prognosis (92.00), life expectancy (86.67), erectile dysfunction (85.00), incontinence (85.00), and irritative urinary symptoms (91.67).

**Table 5.** Micro-averaged F_1_ scores (%) for baseline classifiers and LLM-based prompting strategies across the five communication tradeoffs. The best-performing method for each trade-off is shown in bold.

| Approach | Model/Setting | CP | LE | ED | INC | IUS |
| --- | --- | --- | --- | --- | --- | --- |
| **Baseline** | RoBERTa-large | 42.00 | 73.33 | 41.67 | 53.33 | 78.33 |
|  | PubMedBERT-large | 52.00 | 63.33 | 45.00 | 41.67 | 68.33 |
| **LLM (Rubric)** | Zero-shot | 58.00 | 71.67 | 60.00 | 63.33 | 78.33 |
|  | Few-shot | 76.00 | 86.67 | 73.33 | 65.00 | 85.00 |
| **LLM (Rubric + CoT)** | Zero-shot | 78.00 | 76.67 | 70.00 | 78.33 | 88.33 |
|  | Few-shot | **92.00** | **86.67** | **85.00** | **85.00** | **91.67** |

When comparing baseline classifiers to LLM-based approaches, we observe a substantial performance gap across all tradeoffs. While the fine-tuned transformer models achieved moderate F_1_ scores, most notably RoBERTa-large on IUS and LE (78.33 and 73.33, respectively), they consistently underperformed relative to the prompting-based methods. This discrepancy is likely due to the limited size of the training datasets. With only 35 to 40 training fragments available per tradeoff, the baseline models lacked sufficient data to effectively learn the complex and domain-specific scoring patterns required for this task. In contrast, the use of rubric-based and rubric + CoT prompting strategies enabled LLMs to effectively perform the scoring task with minimal reliance on annotated training data.

Comparing different prompting strategies, we observe two consistent trends. First, incorporating few-shot examples improves performance over zero-shot prompting. For instance, the rubric-only approach improved from 60.00 to 73.33 F_1_ on ED and from 71.67 to 86.67 F_1_ on LE when moving from zero-shot to few-shot prompting. Second, adding CoT reasoning further enhances performance. For example, rubric + CoT with zero-shot prompting achieved 78.00 F_1_ on CP and 88.33 F_1_ on IUS, outperforming rubric-only prompting in both zero-shot and few-shot settings.

## 4. Discussion

Our findings show that rubric + CoT prompting is the most effective strategy for scoring physician communication and the most interpretable. By guiding the LLM to reason through each scoring criterion in a structured, stepwise manner, this approach makes the rationale behind each prediction transparent. In healthcare, such interpretability is critical not only to build trust in AI-assisted evaluations but also to support their integration into clinical practice. Interpretable scoring systems can also offer more actionable and targeted feedback to physicians, highlighting specific aspects of their communication that could be improved.

To examine the performance of this strategy, we conducted a detailed error analysis of the LLM’s predictions using rubric + CoT prompting with few-shot examples. The confusion matrices for each tradeoff (Figure 1) show that, despite strong overall results, the LLM exhibited difficulty distinguishing between adjacent score levels for certain tradeoffs, particularly LE, ED, and INC.

**Fig. 1.**
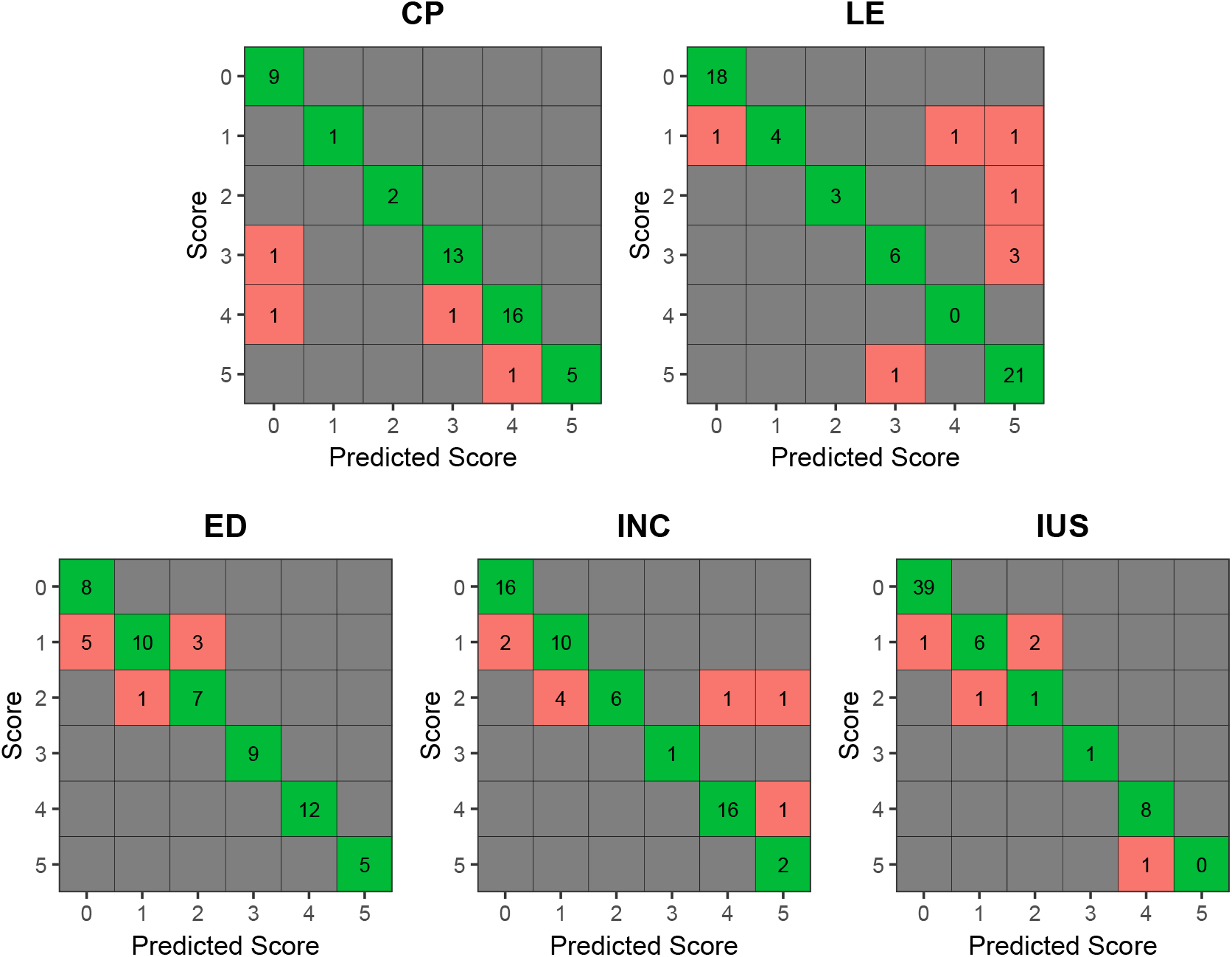
Confusion matrices for each of the five communication tradeoffs using the best-performing system (rubric + CoT with few-shot prompting). Each matrix displays the distribution of predicted versus true scores (0–5).

For LE, the most frequent pattern included three instances where the LLM predicted a score of 5 instead of 3. In these cases, the text provided a rough estimate of years (e.g., “40 years”) alongside general references to the patient’s age (e.g., “you’re 52”). However, such expressions lacked the detailed patient-specific estimate required for score 5.

In the case of ED, five errors involved the LLM predicting score 0 instead of 1. These fragments typically referenced surgical procedures or anatomical structures related to erectile function (e.g., “nerve-sparing techniques”) without explicitly naming ED as a side effect. Although suggestive, the absence of direct mention led the LLM to conclude that the tradeoff was not discussed.

For INC, in four cases the LLM predicted score 1 instead of 2. These cases involved qualitative but vague language, often using generalized statements like “everybody has urinary incontinence”, which the LLM interpreted as a mere mention rather than a qualitative risk estimate.

Importantly, many of the ambiguous cases that challenged the LLM also proved difficult for human annotators to score consistently. These fragments, often characterized by imprecise, indirect, or incomplete references to risk, were a common source of disagreement during the initial annotation phase. Given this context, the performance of our best LLM-based approach is particularly promising: its F_1_ scores are broadly comparable to the pre-adjudication IAA values (see Section 2.2), suggesting that the system can approximate expert-level scoring in this complex classification task.

### 4.1. Limitations and Future Work

While our rubric + CoT prompting with the GPT-4o model achieves strong performance across all five communication tradeoffs, several limitations warrant further investigation. First, the dataset was drawn from a relatively small set of prostate cancer consultations (N=20), which may limit generalizability across institutions, clinicians, and patient populations. Future work will explore larger and more diverse corpora that encompass a wider range of provider communication styles and patient demographics. Second, our approach relies on the accuracy of a preceding NLP-based model used to identify candidate sentences for scoring.^17^ While this model has been previously developed and validated to identify key tradeoff-related sentences in prostate cancer consultations, any misclassifications at this selection stage may impact downstream scoring performance. Future work will explore the integration of these components in real-world clinical workflows to evaluate physician risk communication at scale and ultimately support more informed, patient-centered decision making. Lastly, our methodology has thus far been applied only to prostate cancer, which may limit its applicability to other clinical domains. However, the underlying framework combining scoring rubrics with LLM-based approaches, has the potential to be adapted for risk communication assessment across other cancer types and medical decision-making contexts.

## 5. Conclusion

We present a novel framework for evaluating the quality of physician risk communication in prostate cancer consultations using LLMs. By combining validated scoring rubrics with CoT prompting, our method enables interpretable, sentence-level assessment of how precisely and patient-specifically key tradeoffs are communicated. Across five clinically relevant domains, including cancer prognosis, life expectancy, and three side effects, our best-performing model achieved strong results and expert-level agreement. These findings highlight the potential of LLMs to support scalable, automated evaluations of physician–patient communication and lay the groundwork for real-time feedback tools to improve shared decision-making in oncology and beyond.

## Data Availability

The data used in this study contain protected health information (PHI) and are not publicly available due to privacy and confidentiality regulations.

## Acknowledgments

This research was supported by the National Cancer Institute of the National Institutes of Health under Award Number R01CA290559 to TJD. The content is solely the responsibility of the authors and does not necessarily represent the official views of the National Institutes of Health.

## References

1. T. J. Daskivich, M. S. Litwin and D. F. Penson, Effect of Age, Tumor Risk, and Comorbidity in a U.S. Population–Based Cohort of Men With Prostate Cancer, Annals of Internal Medicine 159, p. 370 (2013).

2. L. F. van de Water, J. J. van Kleef, W. P. M. Dijksterhuis, I. Henselmans, H. G. van den Boorn, N. M. Vaarzon Morel, K. F. Schut, J. G. Daams, E. M. A. Smets and H. W. M. van Laarhoven, Communicating treatment risks and benefits to cancer patients: a systematic review of communication methods, Quality of Life Research 29, 1747 (2020).

3. M. S. Litwin and H.-J. Tan, The Diagnosis and Treatment of Prostate Cancer: A Review, JAMA 317, 2532 (06 2017).

4. T. J. Wilt, T. N. Vo, L. Langsetmo, P. Dahm, T. Wheeler, W. J. Aronson, M. R. Cooperberg, B. C. Taylor and M. K. Brawer, Radical Prostatectomy or Observation for Clinically Localized Prostate Cancer: Extended Follow-up of the Prostate Cancer Intervention Versus Observation Trial (PIVOT), European Urology 77, 713 (2020).

5. D. V. Makarov, K. Chrouser, J. L. Gore, J. Maranchie, M. E. Nielsen, C. Saigal, C. Tessier and Fagerlin, AUA White Paper on Implementation of Shared Decision Making into Urological Practice, Urology Practice 3, 355 (2016).

6. P. C. Albertsen, D. F. Moore, W. Shih, Y. Lin, H. Li and G. L. Lu-Yao, Impact of Comorbidity on Survival Among Men With Localized Prostate Cancer, Journal of Clinical Oncology 29, 1335 (2011).

7. T. J. Daskivich, R. Gale, M. Luu, D. Khodyakov, J. T. Anger, S. J. Freedland and B. Spiegel, Patient Preferences for Communication of Life Expectancy in Prostate Cancer Treatment Consultations, JAMA Surgery 157, 70 (01 2022).

8. A. Bill-Axelson, L. Holmberg, H. Garmo, K. Taari, C. Busch, S. Nordling, M. Häggman, S.-O. Andersson, O. Andrén, G. Steineck, H.-O. Adami and J.-E. Johansson, Radical Prostatectomy or Watchful Waiting in Prostate Cancer — 29-Year Follow-up, New England Journal of Medicine 379, 2319 (2018).

9. R. D. Vromans, M. C. van Eenbergen, G. Geleijnse, S. Pauws, L. V. van de Poll-Franse and E. J. Krahmer, Exploring Cancer Survivor Needs and Preferences for Communicating Personalized Cancer Statistics From Registry Data: Qualitative Multimethod Study, JMIR Cancer 7, p. e25659 (Oct 2021).

10. G. Elwyn, P. J. Barr and S. W. Grande, Patients recording clinical encounters: a path to empowerment? assessment by mixed methods, BMJ Open 5 (2015).

11. T. J. Daskivich, R. Gale, M. Luu, A. Naser-Tavakolian, A. Venkataramana, D. Khodyakov, J. T. Anger, E. Posadas, H. Sandler, B. Spiegel and S. J. Freedland, Variation in Communication of Competing Risks of Mortality in Prostate Cancer Treatment Consultations, The Journal of Urology 208, 301 (2022).

12. T. J. Daskivich, A. Naser-Tavakolian, R. Gale, M. Luu, N. Friedrich, A. Venkataramana, D. Khodyakov, E. Posadas, H. Sandler, B. Spiegel and S. J. Freedland, Variation in communication of side effects in prostate cancer treatment consultations, Prostate Cancer and Prostatic Diseases 28, 145 (2025).

13. T. Daskivich, M. Luu, R. Gale, D. Khodyakov, S. Freedland and B. Spiegel, MP24-03 Development and Validation of a Natural Language Processing System to Identify Information on Key Tradeoffs in Prostate Cancer Treatment Consultations, Journal of Urology 211, p. e392 (2024).

14. A. Hernández Tirado, M. Luu, J. Tan and T. J. Daskivich, MP23-09 A Framework for Evaluating the Quality of Risk Communication in Prostate Cancer Treatment Consultations, Journal of Urology 213, p. e798 (2025).

15. A. Jerfy, O. Selden and R. Balkrishnan, The Growing Impact of Natural Language Processing in Healthcare and Public Health, INQUIRY: The Journal of Health Care Organization, Provision, and Financing 61 (2024).

16. M. Cascella, F. Semeraro, J. Montomoli, V. Bellini, O. Piazza and E. Bignami, The Breakthrough of Large Language Models Release for Medical Applications: 1-Year Timeline and Perspectives, Journal of Medical Systems 48 (2024).

17. R. Zheng, N. A. Friedrich, M. Luu, R. Gale, D. Khodyakov, S. J. Freedland, B. Spiegel and T. J. Daskivich, Development and Validation of a Natural Language Processing System to Assess Quality of Physician Communication in Prostate Cancer Consultations, Prostate Cancer and Prostatic Diseases (2025).

18. J. Wei, X. Wang, D. Schuurmans, M. Bosma, b. ichter, F. Xia, E. Chi, Q.V. Le and D. Zhou, Chain-of-thought prompting elicits reasoning in large language models, in Advances in Neural Information Processing Systems, eds. S. Koyejo, S. Mohamed, A. Agarwal, D. Belgrave, K. Cho and A. Oh (Curran Associates, Inc., 2022).

19. T. Brown, B. Mann, N. Ryder, M. Subbiah, J. D. Kaplan, P. Dhariwal, A. Neelakantan, P. Shyam, G. Sastry, A. Askell, S. Agarwal, A. Herbert-Voss, G. Krueger, T. Henighan, R. Child, A. Ramesh, D. Ziegler, J. Wu, C. Winter, C. Hesse, M. Chen, E. Sigler, M. Litwin, S. Gray, B. Chess, J. Clark, C. Berner, S. McCandlish, A. Radford, I. Sutskever and D. Amodei, Language Models are Few-Shot Learners, in Advances in Neural Information Processing Systems, eds. H. Larochelle, M. Ranzato, R. Hadsell, M. Balcan and H. Lin (Curran Associates, Inc., 2020).

20. M. L. McHugh, Interrater reliability: the kappa statistic, Biochemia Medica 22, 276 (2012).

21. T. Kojima, S. S. Gu, M. Reid, Y. Matsuo and Y. Iwasawa, Large language models are zero-shot reasoners, in Advances in Neural Information Processing Systems, eds. S. Koyejo, S. Mohamed, Agarwal, D. Belgrave, K. Cho and A. Oh (Curran Associates, Inc., 2022).

22. Y. Ge, Y. Guo, S. Das, M. A. Al-Garadi and A. Sarker, Few-shot learning for medical text: A review of advances, trends, and opportunities, Journal of Biomedical Informatics 144, p. 104458 (2023).

23. J. Devlin, M.-W. Chang, K. Lee and K. Toutanova, BERT: Pre-training of Deep Bidirectional Transformers for Language Understanding, in Proceedings of the 2019 Conference of the North American Chapter of the Association for Computational Linguistics: Human Language Technologies, Volume 1 (Long and Short Papers), (Association for Computational Linguistics, Minneapolis, Minnesota, June 2019).

24. A. Vaswani, N. Shazeer, N. Parmar, J. Uszkoreit, L. Jones, A. N. Gomez, L. u. Kaiser and I. Polosukhin, Attention is All you Need, in Advances in Neural Information Processing Systems, eds. I. Guyon, U. V. Luxburg, S. Bengio, H. Wallach, R. Fergus, S. Vishwanathan and R. Garnett (Curran Associates, Inc., 2017).

25. Y. Liu, M. Ott, N. Goyal, J. Du, M. Joshi, D. Chen, O. Levy, M. Lewis, L. Zettlemoyer and V. Stoyanov, RoBERTa: A Robustly Optimized BERT Pretraining Approach (2019).

26. R. Tinn, H. Cheng, Y. Gu, N. Usuyama, X. Liu, T. Naumann, J. Gao and H. Poon, Fine-tuning large neural language models for biomedical natural language processing, Patterns 4, p. 100729 (2023).

